# Association between physical-activity trajectories and cognitive decline in adults 50 years of age or older

**DOI:** 10.1101/2021.04.13.21255381

**Authors:** Boris Cheval, Zsófia Csajbók, Tomáš Formánek, Stefan Sieber, Matthieu P. Boisgontier, Stéphane Cullati, Pavla Cermakova

## Abstract

**Objectives:** To investigate the associations of physical-activity trajectories with the level of cognitive performance and its decline in adults 50 years of age or older.

**Methods:** We studied 38729 individuals (63 ± 9 years; 57% women) enrolled in the Survey of Health, Ageing and Retirement in Europe (SHARE). Physical activity was self-reported and cognitive performance was assessed based on immediate recall, verbal fluency, and delayed recall. Physical-activity trajectories were estimated using growth mixture modelling and linear mixed effects models were used to investigate the associations between the trajectories and cognitive performance.

**Results:** The models identified two physical-activity trajectories of physical activity: constantly-high physical activity (N=27634: 71%) and decreasing physical activity (N=11095; 29%). Results showed that participants in the decreasing physical-activity group exhibited a lower level of cognitive performance compared to the high physical-activity group (immediate recall: ß=0.94; 95% confidence interval [CI]=0.92 to 0.95; verbal fluency: ß=0.98; 95% CI=0.97 to 0.98; delayed recall: ß=0.95; 95% CI=0.94 to 0.97). Moreover, compared with participants in the constantly-high physical-activity group, participants in the decreasing physical-activity group showed a steeper decline in all cognitive measures (immediate recall: ß=-0.04; 95% CI=-0.05 to −0.04; verbal fluency: ß=-0.22; 95% CI=-0.24 to −0.21; delayed recall: ß=-0.04; 95% CI=-0.05 to −0.04).

**Conclusions:** Physical-activity trajectories are associated with the level and evolution of cognitive performance in adults over 50 years. Specifically, our findings suggest that a decline in physical activity over multiple years is associated with a lower level and a steeper decline in cognitive performance.

## INTRODUCTION

Physical activity (PA) and cognitive performance (CP) are strongly linked and have been shown to decline with ageing.^1-5^ Multiple observational and interventional studies have demonstrated that a higher level of PA is associated with better CP in old age.^6-14^ However, most of these studies focused on the association between PA levels and CP. Yet, disregarding the intra-individual evolution of PA over time may have bias the observed association with CP.^15^ Because PA is a complex behavior that evolves over time,^15^ examining the associations between life-course PA trajectories and CP is warranted.

Recent studies have attempted to address this gap by considering the evolution of PA over time.^16-25^ However, most of those studies were based on conventional methods to identify groups with different PA trajectories, which typically use clinical or empirical cut-points.^16-21^ To reduce the reliance on these cut-points, data-driven approaches such as growth mixture modelling have been proposed.^26, 27^ These models do not require a priori trajectory classifications as the trajectories emerge from the data.^22, 28^ Using this approach, studies have shown PA trajectories in older adults^29, 30^ were associated with multiple health outcomes including disability, cardiovascular diseases, and all-cause mortality.^22-25, 31^ However, the associations of PA trajectories with CP and is unknown.

In this study, we aimed to investigate the associations of PA trajectories with CP level and its decline in adults 50 years of age or older. We hypothesized that unfavorable profiles (e.g., individuals with decreasing PA over time) would be associated with lower level and steeper decline of CP compared to more favorable profiles (e.g., individuals maintaining high PA levels).

## METHODS

### Design and population

Data were drawn from the Survey of Health, Ageing and Retirement in Europe (SHARE), an ongoing population-based study of health, social network, and economic conditions of community-dwelling middle-aged and older individuals, living in 28 European countries and Israel (N=139556). SHARE was described in detail elsewhere.^32^ Briefly, participants were sampled based on probability selection methods. Individuals eligible for the study were people aged 50 years or older and their partners, irrespective of age. Data was collected using computer-assisted personal interviewing (CAPI) in participants’ homes. The study was initiated in 2004 and followed by six subsequent waves with approximately two-year intervals and wave 7 being completed in 2017. Wave 3 was devoted to data collection related to childhood histories (SHARELIFE). This wave did not contain any data related to PA and CP and was therefore not used in the current study. This study was carried out in accordance with the Declaration of Helsinki. SHARE has been approved by the Ethics Committee of the University of Mannheim (waves 1-4) and the Ethics Council of the Max Plank Society (waves 4-7). All participants provided a written informed consent. Data were pseudo-anonymized and participants were informed about the storage and use of the data and their right to withdraw consent.

In the present study, we included a total of 38729 participants who fulfilled the following criteria: 1) age = 50 years or older at baseline, 2) measures of PA in at least three waves, 3) measures of cognition in at least two waves, and 4) no report of diagnosed dementia (Figure 1).

**Figure 1.**
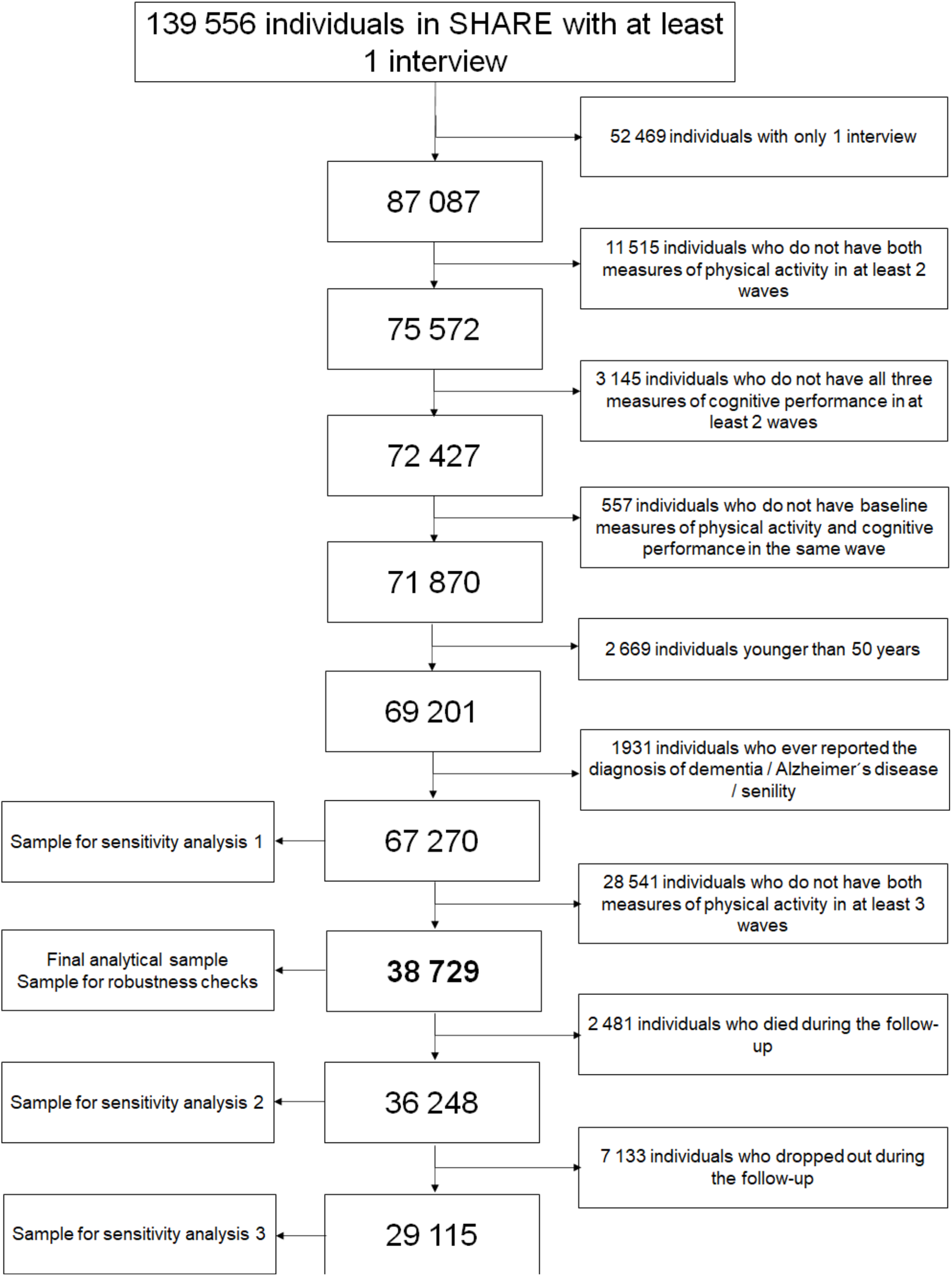
Selection of the study sample

### Measures

#### Physical activity

PA was assessed as part of CAPI in waves 1, 2, 4, 5, 6, and 7 using two questions. “*How often do you engage in vigorous physical activity, such as sports, heavy housework, or a job that involves physical labor?”* and “*How often do you engage in activities that require a low or moderate level of energy such as gardening, cleaning the car, or doing a walk?”* Participants answered on a four-point scale (0=*Hardly ever, or never*; 1=*One to three times a month*; 2=*Once a week*; 3=*More than once a week*). An overall score of PA ranging from 0 to 6 was created by summing up the scores on the two questions, with higher scores reflecting greater PA.

#### Cognitive tests

CP was assessed in wave 1, 2, 4, 5, 6, and 7 using validated tests of verbal fluency, immediate recall, and delayed recall. In the verbal fluency test,^33^ participants were instructed to name as many different animals as they could think of in one minute. The score was the total number of correctly named animals, with a higher score indicating higher verbal fluency. Immediate and delayed recall were assessed using an adapted 10-word delayed-recall test.^34^ In the immediate-recall test, participants first listened to a 10-word list that was read out loud by the interviewer. Then, immediately after the reading of this list, participants were asked to recall as many words as possible. At the end of the cognitive testing session, the participants were asked again to recall any of the words from the list, which captured delayed recall score. Both scores ranged from 0 to 10, with a higher score indicating greater performance.

### Covariates

Models were adjusted for sociodemographic and health-related characteristics identified as potentially confounding and mediating factors in the association between PA and CP level and decline were identified.^35-39^ The value of the covariates was the one at baseline if all PA and CP data were available. In case of missing data at baseline, the value of the covariate was taken from the closest available wave. The selected sociodemographic factors were area of Europe (Western Europe, Scandinavia, Southern Europe, Central and Eastern Europe), age (years), sex (male, female), education (seven categories based on the International Standard Classification of Education 1997),^40^ residence (big city, suburbs or outskirts of a big city, large town, small town, rural area or village), household size (number of members), partner in household (yes, no), household net worth (standardized difference between household gross financial assets and financial liabilities), current job situation (working, not working), number of children, number of grandchildren, and attrition (non-participation in all the waves or death during the follow-up; yes, no). Health-related characteristics were the number of limitations in instrumental activities of daily living (IADL), number of depressive symptoms assessed with the EURO-D scale,^41^ number of chronic diseases, body mass index (continuous variable), mobility limitations index (number of limitations), smoking (ever smoked daily, never smoked daily), alcohol use (more than 2 glasses of alcohol almost every day, less), frequency of eating fruits and vegetables (every day, 3-6 times a week, twice a week, once a week, less than once a week).

### Statistical analyses

#### PA trajectories

As in our previous study on trajectories of depressive symptoms,^28^ growth mixture modelling with maximum likelihood estimation was used to identify latent trajectories of PA.^42^ This approach estimates latent classes following similar trajectories of PA over time with a high probability. Consistent with previous guidelines and literature,^28, 43^ we first freely estimated time slopes in a latent basis growth model that was entered to the growth mixture model (i.e., the classification). Second, the most parsimonious model among those with different number of PA trajectories (i.e., classes) was determined using the following indicators: Akaike Information Criterion, Bayesian Information Criterion, sample-size adjusted Bayesian information criterion, Vuong-Lo-Mendell-Rubin likelihood ratio test, Luo-Mendell-Rubin adjusted likelihood ratio test, and bootstrap likelihood ratio test (see Supplementary Methods). Finally, we checked the number of individuals within each PA trajectory group to ensure an adequate sample size.

#### Association between PA trajectories and baseline characteristics

First, independent samples t-tests and χ^2^ tests were used to compare the differences in baseline characteristics between PA trajectories. Second, a multivariable analysis using logistic regression was performed to estimate odds ratio (OR) with 95% confidence interval (CI) for the association between participants’
s baseline characteristics and PA trajectories.

#### Association between PA trajectories and the level and rate of decline of CP

We used linear mixed effects models to investigate the associations of PA trajectories with the level of CP and its rate of decline over time. The models included time in years since baseline, PA trajectory, their interaction term (PA trajectory × time), and covariates. Model 1 was adjusted for age and sex. Other sociodemographic covariates (birth cohort, region, education, residence, household size, partner in household, household net worth, current job situation, number of children, number of grandchildren and attrition) were included in Model 2. Health-related characteristics (limitations in instrumental activities of daily living, depressive symptoms, number of chronic diseases, body mass index, mobility limitations index, smoking, alcohol use and eating behavior) were included in Model 3. We adjusted for covariates group-wise in three steps to assess whether sociodemographic and health-related characteristics may explain the observed associations. The model random structure encompassed random intercepts for participants and random linear slopes for time. In addition, we stratified the dataset according PA trajectories, and (1) fitted a crude model containing only time and (2) Models 1-3 (except of interaction term) for each cognitive test, separately. Based on the fit of the crude stratified model, we visualized the cognitive decline across trajectories of PA and per cognitive tests.

#### Sensitivity and robustness analyses

We performed three sensitivity analyses (SA), in which we replicated the growth mixture modelling on (1) a sample of participants having at least two measures of PA, two measures of CP, and no diagnosis of dementia (SA1: N = 67270), (2) a sample only including surviving participants (SA2: N = 36248), and (3) a sample of participants who neither dropped out nor died during the follow-up of the study (SA3: N = 29115). We conducted two robustness analyses, in which we performed the growth mixture modelling on (a) the moderate physical activity measure only and (b) on the vigorous physical activity measure only.

### Data availability

Access to the SHARE data is provided free of charge on the basis of a release policy that gives quick and convenient access to all scientific users worldwide after individual registration. All details about the application and registration process can be found on this website: http://www.share-project.org. The study protocol and syntax of the statistical analysis will be shared upon request from the corresponding author of this study.

## RESULTS

### Physical-activity trajectories

The study sample included 38729 adults 50 years of age or older (63 years on average, 57% women). Two PA trajectory groups emerged from the growth mixture modelling as the best model for describing the longitudinal data: constantly-high PA (N=27634: 71%) and decreasing PA (N=11095; 29%) (Figure 2). This two-trajectory solution showed acceptable fit to the data, although with relatively low entropy (0.603), thereby indicating some overlap between the two classes. Results from the process of model selection, including the results of the sensitivity and robustness analyses, are provided in the supplementary materials.

**Figure 2.**
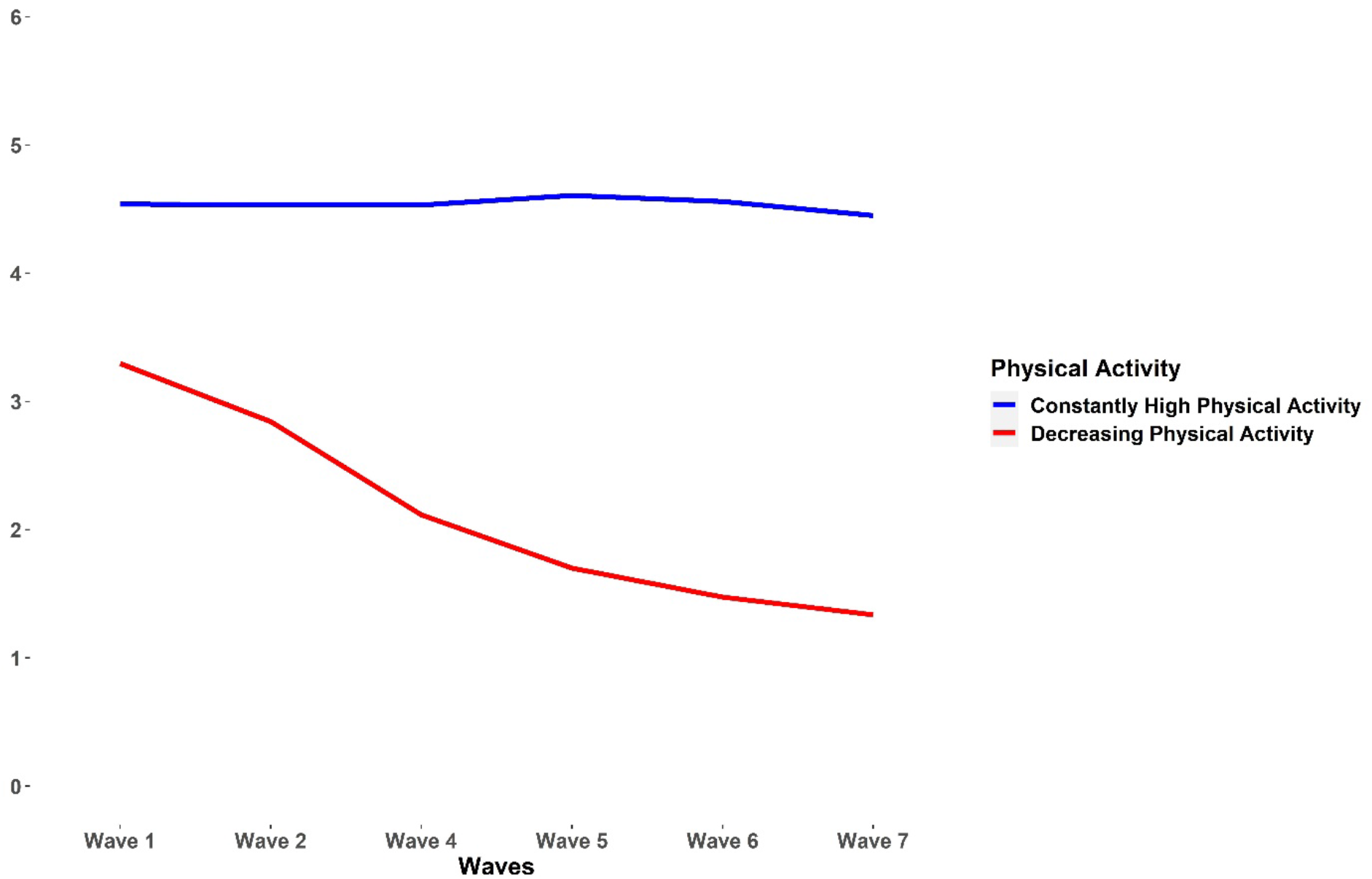
Physical activity trajectories

### Physical-activity trajectories and baseline characteristics of the participants

Table 1 summarizes the characteristics of the participants stratified by PA trajectory. Compared with the constantly-high PA group, individuals with decreasing PA showed lower baseline level in the three measures of CP (mean verbal fluency 17.9 vs. 21.5; mean immediate recall 4.7 vs. 5.5; mean delayed recall 3.2 vs. 4.1; *p*s<0.001). In addition, individuals with decreasing PA were older (p<0.001) and predominantly women (p<0.001).

**Table 1.**
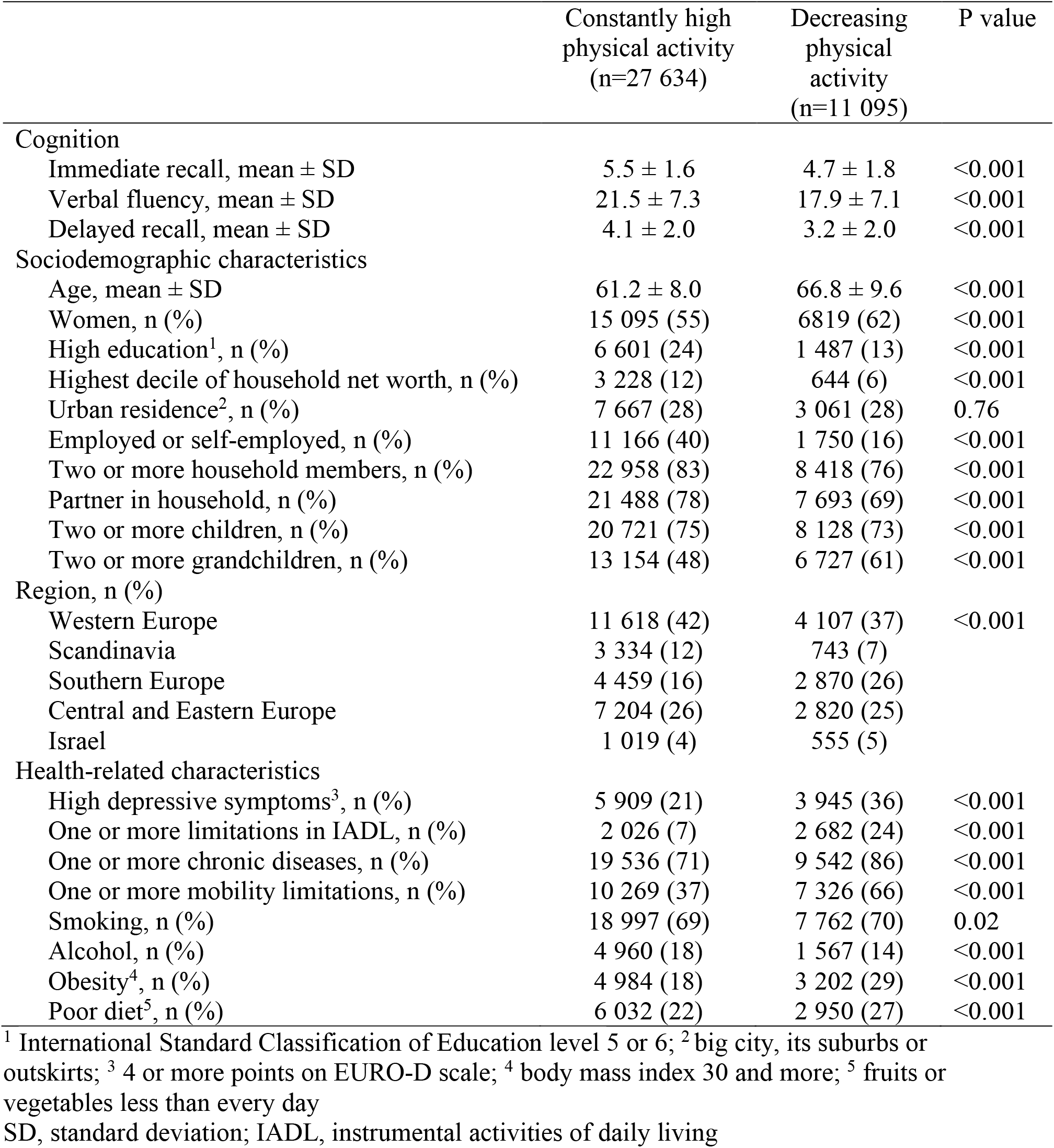
Baseline characteristics of the participants across PA trajectories

|  | Constantly high<br>physical activity<br>(n=27 634) | Decreasing<br>physical<br>activity<br>(n=11 095) | P value |
| --- | --- | --- | --- |
| Cognition |  |  |  |
| Immediate recall, mean ± SD | 5.5 ± 1.6 | 4.7 ± 1.8 | <0.001 |
| Verbal fluency, mean ± SD | 21.5 ± 7.3 | 17.9 ± 7.1 | <0.001 |
| Delayed recall, mean ± SD | 4.1 ± 2.0 | 3.2 ± 2.0 | <0.001 |
| Sociodemographic characteristics |  |  |  |
| Age, mean ± SD | 61.2 ± 8.0 | 66.8 ± 9.6 | <0.001 |
| Women, n (%) | 15 095 (55) | 6819 (62) | <0.001 |
| High education<sup>1</sup>, n (%) | 6 601 (24) | 1 487 (13) | <0.001 |
| Highest decile of household net worth, n (%) | 3 228 (12) | 644 (6) | <0.001 |
| Urban residence<sup>2</sup>, n (%) | 7 667 (28) | 3 061 (28) | 0.76 |
| Employed or self-employed, n (%) | 11 166 (40) | 1 750 (16) | <0.001 |
| Two or more household members, n (%) | 22 958 (83) | 8 418 (76) | <0.001 |
| Partner in household, n (%) | 21 488 (78) | 7 693 (69) | <0.001 |
| Two or more children, n (%) | 20 721 (75) | 8 128 (73) | <0.001 |
| Two or more grandchildren, n (%) | 13 154 (48) | 6 727 (61) | <0.001 |
| Region, n (%) |  |  |  |
| Western Europe | 11 618 (42) | 4 107 (37) | <0.001 |
| Scandinavia | 3 334 (12) | 743 (7) |  |
| Southern Europe | 4 459 (16) | 2 870 (26) |  |
| Central and Eastern Europe | 7 204 (26) | 2 820 (25) |  |
| Israel | 1 019 (4) | 555 (5) |  |
| Health-related characteristics |  |  |  |
| High depressive symptoms<sup>3</sup>, n (%) | 5 909 (21) | 3 945 (36) | <0.001 |
| One or more limitations in IADL, n (%) | 2 026 (7) | 2 682 (24) | <0.001 |
| One or more chronic diseases, n (%) | 19 536 (71) | 9 542 (86) | <0.001 |
| One or more mobility limitations, n (%) | 10 269 (37) | 7 326 (66) | <0.001 |
| Smoking, n (%) | 18 997 (69) | 7 762 (70) | 0.02 |
| Alcohol, n (%) | 4 960 (18) | 1 567 (14) | <0.001 |
| Obesity<sup>4</sup>, n (%) | 4 984 (18) | 3 202 (29) | <0.001 |
| Poor diet<sup>5</sup>, n (%) | 6 032 (22) | 2 950 (27) | <0.001 |

<sup>1</sup> International Standard Classification of Education level 5 or 6; <sup>2</sup> big city, its suburbs or outskirts; <sup>3</sup> 4 or more points on EURO-D scale; <sup>4</sup> body mass index 30 and more; <sup>5</sup> fruits or vegetables less than every day
SD, standard deviation; IADL, instrumental activities of daily living

Table 2 shows the results of the logistic regression models testing the association between PA trajectories and participants’ sociodemographic and health-related characteristics. Compared to individuals with the constantly-high PA, decreasing PA was associated with older age, lower education, lower household wealth, non-urban residence, not working, living without a partner, greater depressive symptoms, more limitations in IADL and mobility, more chronic diseases, smoking, obesity and poor diet. When compared to Western Europe, decreasing PA was more likely to occur in Southern Europe and Israel and less likely in Scandinavia and Central and Eastern Europe.

**Table 2.**
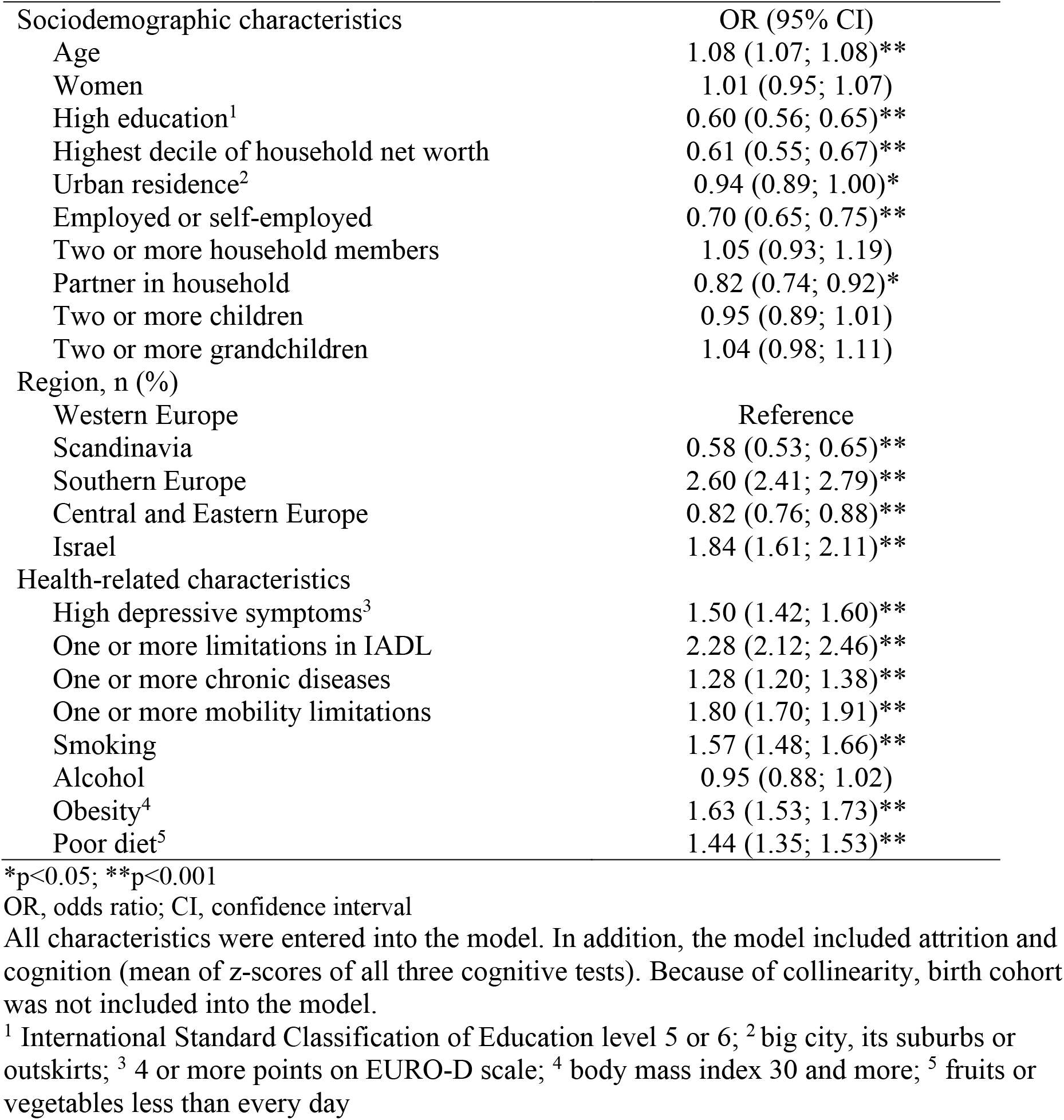
Associations of participants’
s characteristics with decreasing physical activity

### Physical-activity trajectories and cognitive performance

Table 3 shows the results of the linear mixed effects models. Compared with individuals from the constantly high-PA group, individuals in the decreasing PA group had a significantly lower level of baseline CP, independently of all covariates (p<0.001 for all three measures in Model 1, 2 and 3). Individuals in the decreasing PA group also had a steeper decline of CP in all CP measures, independently from covariates, as showed by the PA trajectory × time interaction (p<0.001 for all three measures in Models 1, 2, and 3).

**Table 3.**
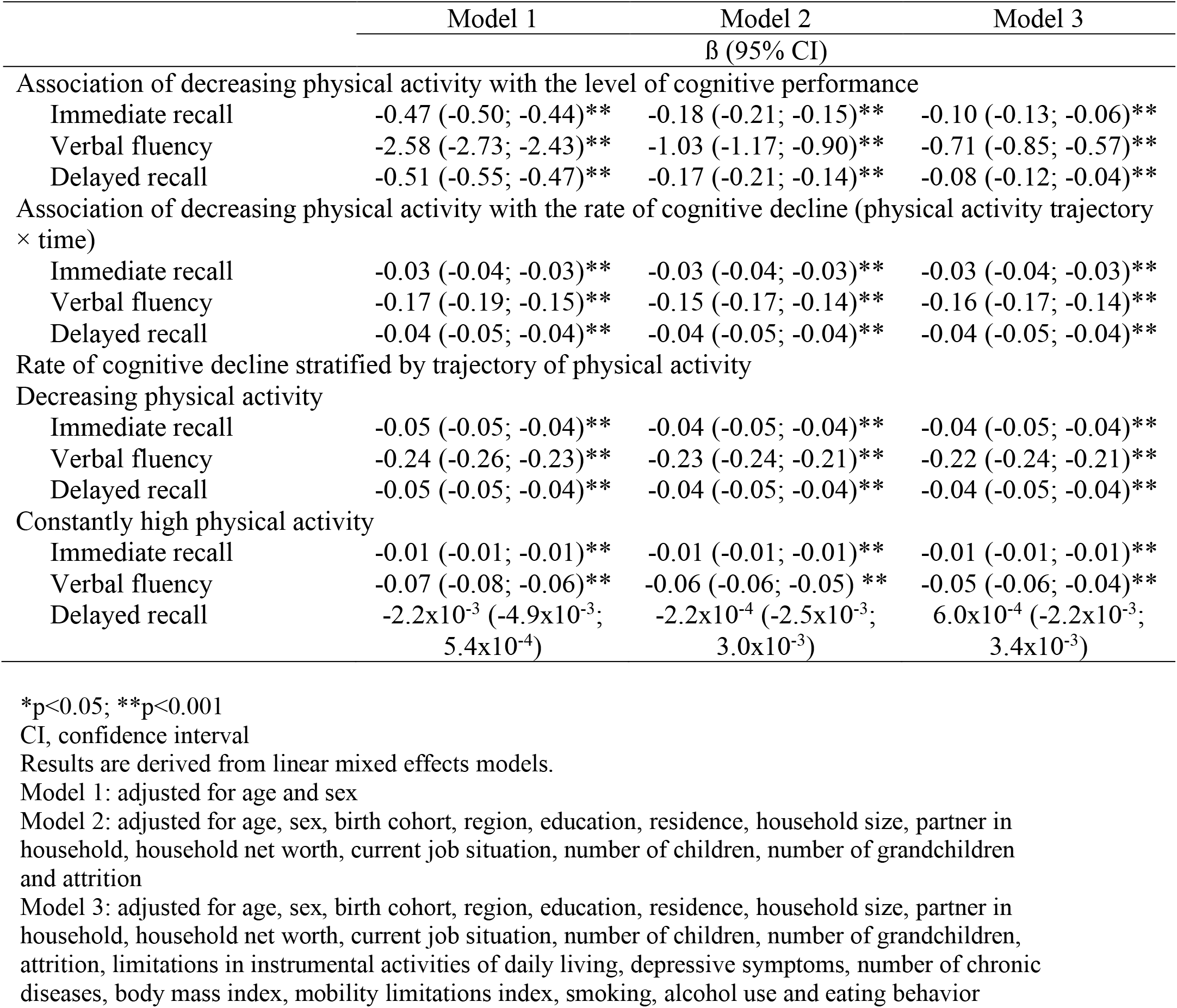
Level of cognitive performance and rate of cognitive decline per year across trajectories of physical activity

|  | Model 1 | Model 2 | Model 3 |
| --- | --- | --- | --- |
| | $\beta$ (95% CI) | | |
| Association of decreasing physical activity with the level of cognitive performance |  |  |  |
| Immediate recall | -0.47 (-0.50; -0.44)** | -0.18 (-0.21; -0.15)** | -0.10 (-0.13; -0.06)** |
| Verbal fluency | -2.58 (-2.73; -2.43)** | -1.03 (-1.17; -0.90)** | -0.71 (-0.85; -0.57)** |
| Delayed recall | -0.51 (-0.55; -0.47)** | -0.17 (-0.21; -0.14)** | -0.08 (-0.12; -0.04)** |
| Association of decreasing physical activity with the rate of cognitive decline (physical activity trajectory × time) | | | |
| Immediate recall | -0.03 (-0.04; -0.03)** | -0.03 (-0.04; -0.03)** | -0.03 (-0.04; -0.03)** |
| Verbal fluency | -0.17 (-0.19; -0.15)** | -0.15 (-0.17; -0.14)** | -0.16 (-0.17; -0.14)** |
| Delayed recall | -0.04 (-0.05; -0.04)** | -0.04 (-0.05; -0.04)** | -0.04 (-0.05; -0.04)** |
| Rate of cognitive decline stratified by trajectory of physical activity |  |  |  |
| Decreasing physical activity |  |  |  |
| Immediate recall | -0.05 (-0.05; -0.04)** | -0.04 (-0.05; -0.04)** | -0.04 (-0.05; -0.04)** |
| Verbal fluency | -0.24 (-0.26; -0.23)** | -0.23 (-0.24; -0.21)** | -0.22 (-0.24; -0.21)** |
| Delayed recall | -0.05 (-0.05; -0.04)** | -0.04 (-0.05; -0.04)** | -0.04 (-0.05; -0.04)** |
| Constantly high physical activity |  |  |  |
| Immediate recall | -0.01 (-0.01; -0.01)** | -0.01 (-0.01; -0.01)** | -0.01 (-0.01; -0.01)** |
| Verbal fluency | -0.07 (-0.08; -0.06)** | -0.06 (-0.06; -0.05)** | -0.05 (-0.06; -0.04)** |
| Delayed recall | -2.2x10<sup>-3</sup> (-4.9x10<sup>-3</sup>; 5.4x10<sup>-4</sup>) | -2.2x10<sup>-4</sup> (-2.5x10<sup>-3</sup>; 3.0x10<sup>-3</sup>) | 6.0x10<sup>-4</sup> (-2.2x10<sup>-3</sup>; 3.4x10<sup>-3</sup>) |

\* $p < 0.05$ ; \*\* $p < 0.001$
CI, confidence interval
Results are derived from linear mixed effects models.
Model 1: adjusted for age and sex
Model 2: adjusted for age, sex, birth cohort, region, education, residence, household size, partner in household, household net worth, current job situation, number of children, number of grandchildren and attrition

When stratified by the PA trajectory, individuals from the constantly-high PA group had negligible rates of decline in immediate recall, verbal fluency and none in delayed recall, while individuals from the decreasing PA group showed steeper rates of decline in all three cognitive measures (Figure 3). Adjustment for covariates only slightly attenuated the rates of decline. In the fully-adjusted models, individuals from the decreasing PA group had a steeper decline in all cognitive measures (immediate recall: ß=-0.04; 95% CI=-0.05 to −0.04; verbal fluency: ß=-0.22; 95% CI=-0.24 to −0.21; delayed recall: ß=-0.04; 95% CI=-0.05 to −0.04), in comparison to the other group. In contrast, individuals from the constantly high PA group only demonstrated a small decline in immediate recall (ß −0.01; 95% CI −0.01 to −0.01) and verbal fluency (ß=-0.05; 95% CI −0.06 to −0.04) and none in delayed recall (ß=6.0×10^−4^; 95% CI −2.2×10^−3^ to 3.4×10^−3^).

**Figure 3.**
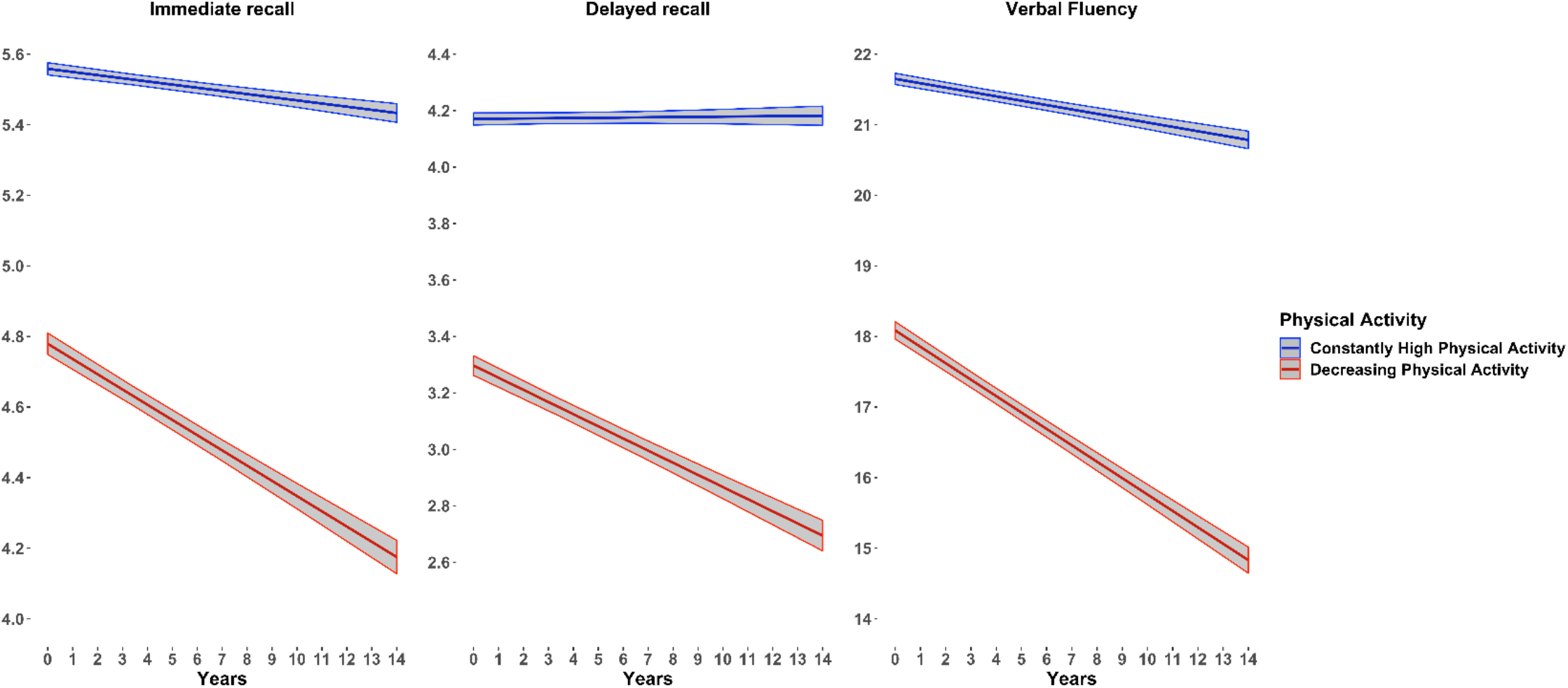
Crude yearly rates of cognitive decline across trajectories of physical activity

#### Sensitivity and robustness analyses

The SA and robustness analyses (Supplementary tables S4, S5, and S6) yielded similar results as the main analysis. Specifically, we replicated the 2-class solution (see supplementary methods). Furthermore, the linear mixed effects models showed that, compared with individuals from the constantly high PA group, individuals from the decreasing PA group had a significantly lower level of baseline in the three CP measures and exhibited a steeper decline of those measures. The only difference is that the associations between the PA groups and the decline of immediate and delayed recall over time did not remain significant after the adjustment for all the covariates.

## DISCUSSION

In the present large-scale longitudinal multicentric study of adults aged 50 years and older, we identified two distinct PA trajectories: constantly-high PA and decreasing PA. These profiles of PA trajectories were associated with the level of CP and the rate of its decline over time. Compared with the constantly-high PA profile, the decreasing PA profile was significantly associated with lower baseline levels of CP, and with a significantly steeper decline over time of CP. These associations were only slightly attenuated after adjustment for the sociodemographic and health-related characteristics. These findings suggest that the CP decline over time is mirrored in the longitudinal changes of PA.

Our results complement the literature on the association of PA and CP during ageing from a longitudinal perspective. Using growth mixture modelling, we identified two distinct PA trajectories over time. One PA trajectory showed PA levels that remained constant across time, a finding that is consistent with previous studies that observed stable PA trajectories in old age.^22, 23, 44^ The second PA trajectory that emerged showed a decreased PA across time. This result is in line with previous studies that have observed a reduction in PA at old age.^25, 45^ However, in addition to these two PA trajectories, other studies found increasing PA trajectories in middle aged and older adults.^24, 45^ Moreover, studies often reported three or four PA trajectory groups, which contrasts with the two PA trajectories that have emerged from the current data.^45^ This discrepancy may be explained by the features of the scale that was used to measure PA (i.e., 2-items measuring the frequency of usual moderate and vigorous PA), which was associated with low variance. As such, it was difficult to identify clearly distinct PA trajectories as indicated by the low level of entropy observed between our two selected PA groups.

In the present study, 71% of our sample were classified in the more favorable profile (i.e., constantly high PA), while only 29% were classified in the unfavorable profile (i.e., decreasing PA). This result is rather consistent with previous studies observing that about 30% of older adults showed a decreasing PA trajectory across time.^22, 24, 25^ However, while this finding is very encouraging in terms of public health, it must be interpreted with caution given that, in our study, PA was assessed using a self-reported questionnaire, which may have biased the estimation of participants’ PA behaviors. Furthermore, when age is used as the time scale instead of the wave of measurement (i.e., using an accelerated longitudinal design assessing PA evolution from 65 to 100 years), results showed a clear decline in PA across aging in all the PA groups.^23, 31^

We observed that a number of sociodemographic and health-related characteristics are associated with the PA trajectories. In particular, older age, worse socioeconomic status, poor lifestyle profile and worse health were associated with increased odds of belonging to the unfavorable PA group. Our findings are consistent with previous studies that have investigated the multiple correlates of higher engagement in PA.^35, 39^ For example, chronic conditions,^36^ depressive symptoms,^37^ or lower level education,^38^ have been found to be associated with a lower level of engagement in PA. Our results confirm these associations but also reveal that these factors are not only associated with the level of PA engagement, but also with its evolution across time. Even though causality cannot be established due to observational design of this study, these findings indicate that decline in PA and CP could be predicted. Focusing health care and preventive efforts on subgroups of individuals with a low socioeconomic status and poor health profile could contribute to prevention of decreasing PA and CP.

The strong relationship between higher levels of PA and better cognitive function are well-established,^1-5^ and our results confirm this association. To the best of our knowledge, our study was the first to investigate this association using a data-driven approach to identify PA trajectories. One study has estimated trajectories of PA over 28 years in people with or without dementia and found that PA started to decline nine years before the diagnosis of dementia.^46^ A result that rather suggests that changes in PA may simply result from the decline in CP, which is consistent with other studies that have observed an association from changes in CP to changes in PA.^1, 47^ Moreover, intervention studies in older adults observed a protective effect of an increased PA on CP,^12, 14^ although other studies did not.^48-50^ However, these studies focused on relatively short-term changes in PA (i.e., from one time of measurement to another one; or before and after an intervention), but disregarded the long-term trajectories of PA across time. Overall, our results are in line with previous studies showing that changes in PA are associated with changes in CP.

Several mechanisms have been suggested to explain the association between higher level of PA and a maintained level of CP.^14, 51-57^ For example, PA has been associated with increased brain plasticity, angiogenesis, synaptogenesis, and neurogenesis primarily through the release of growth factors such as brain-derived neurotrophic factor, insulin-like growth factor-1 and vascular endothelial growth factor. ^53, 54, 56^ Moreover, the cognitive demands inherent in many types of PA, which may include planning, reasoning, decision-making, and multitasking, may also have positive effects on the brain.^57-60^ Alternatively, additional mechanisms have been suggested to explain the association in the other direction – i.e., from cognitive functioning to higher engagement in physical activity.^61-64^ In particular, anchored within the theory of effort minimization in physical activity (TEMPA),^65^ these studies showed that cognitive function are critical to counteract the automatic tendency to effort minimization, thereby favoring engagement in PA.

Among the strengths of the present study are the large sample size, the longitudinal design, the data driven approach to identify PA trajectories across time, and the reliance on three measures of CP based on established procedures. However, some limitations should be noted. First, PA was assessed using a self-reported questionnaire, which may have reduced measurement validity. Second, the two PA groups differed in both the initial PA levels (i.e., high vs. moderate) and the evolution of PA across time (i.e., maintaining vs. decreasing). As such, it was not possible to disentangle the influence of these two factors (i.e., level or slope) on CP. Future research should rely on a more detailed and reliable questionnaire or on device-based measures of PA to be able to better discriminate between PA trajectories, and of their associations with CP. Likewise, as the PA groups differed regarding overall health related condition, we cannot exclude that the a steeper cognitive decline observed the unfavorable PA profile can result from this poorer health condition rather than the change in PA. Furthermore, as it cannot be avoided in long-term longitudinal studies, we cannot exclude a selection bias due to attrition. Yet, to minimize this bias, our statistical analyses were adjusted for attrition and we conducted sensitivity analyses excluding participants who dropped out or died during the follow-up. Finally, the correlational nature of the SHARE design cannot guarantee causal links between PA trajectories and CP decline.

In conclusion, these findings confirm the hypothesis that an unfavorable PA profile may be associated with weaker CP in old age, thereby supporting the need to promote effective strategies to help individuals to maintain their PA levels over the life course.

## Data Availability

This SHARE dataset is available at http://www.share-project.org/data-access.html.

## Acknowledgements

*This paper uses data from SHARE Waves 1, 2, 3 (SHARELIFE), 4, 5,6, 7 and 8 (DOIs: <u>10..6103/SHARE.w1.600</u>, <u>10..6103/SHARE.w2.600</u>, <u>10..6103/SHARE.w3.600</u>, <u>10..6103/SHARE.w4.600</u>, <u>10..6103/SHARE.w5.600</u>, <u>10..6103/SHARE.w6.600, 10.6103/SHARE.w7.711, 10.6103/SHARE.w8cabeta.001</u>)*. The SHARE data collection was primarily funded by the European Commission through FP5 (QLK6-CT-2001-00360), FP6 (SHARE-I3: RII-CT-2006-062193, COMPARE: CIT5-CT-2005-028857, SHARELIFE: CIT4-CT-2006-028812) and FP7 (SHARE-PREP: no.211909, SHARE-LEAP: no.227822, SHARE M4: no.261982). Additional funding from the German Ministry of Education and Research, the Max Planck Society for the Advancement of Science, the U.S. National Institute on Aging (U01_AG09740-13S2, P01_AG005842, P01_AG08291, P30_AG12815, R21_AG025169, Y1-AG-4553-01, IAG_BSR06-11, OGHA_04-064, HHSN271201300071C) and from various national funding sources is gratefully acknowledged (see www.share-project.org).

## References

1. Cheval B, Orsholits D, Sieber S, Courvoisier DC, Cullati S, Boisgontier MP. Relationship between decline in cognitive resources and physical activity. Health Psychol 2020;39:519–528.

2. Cheval B, Sieber S, Guessous I, et al. Effect of early-and adult-life socioeconomic circumstances on physical inactivity. Med Sci Sports Exerc 2018;50:476–485.

3. DiPietro L. Physical activity in aging: changes in patterns and their relationship to health and function. The Journals of Gerontology Series A: Biological Sciences and Medical Sciences 2001;56:13–22.

4. Levy R. Aging-associated cognitive decline. Int Psychogeriatr 1994;6:63–68.

5. Sebastiani P, Andersen SL, Sweigart B, et al. Patterns of multi-domain cognitive aging in participants of the Long Life Family Study. Geroscience 2020;42:1335–1350.

6. Baumgart M, Snyder HM, Carrillo MC, Fazio S, Kim H, Johns H. Summary of the evidence on modifiable risk factors for cognitive decline and dementia: a population-based perspective. Alzheimer Dement 2015;11:718–726.

7. Sofi F, Valecchi D, Bacci D, et al. Physical activity and risk of cognitive decline: a meta-analysis of prospective studies. J Intern Med 2011;269:107–117.

8. Morgan GS, Gallacher J, Bayer A, Fish M, Ebrahim S, Ben-Shlomo Y. Physical activity in middle-age and dementia in later life: findings from a prospective cohort of men in Caerphilly, South Wales and a meta-analysis. J Alzheimers Dis 2012;31:569–580.

9. Blondell SJ, Hammersley-Mather R, Veerman JL. Does physical activity prevent cognitive decline and dementia?: A systematic review and meta-analysis of longitudinal studies. BMC Public Health 2014;14:510.

10. Hamer M, Terrera GM, Demakakos P. Physical activity and trajectories in cognitive function: English Longitudinal Study of Ageing. J Epidemiol Community Health 2018;72:477–483.

11. Lindwall M, Cimino CR, Gibbons LE, et al. Dynamic associations of change in physical activity and change in cognitive function: Coordinated analyses of four longitudinal studies. J Aging Res 2012;2012:793598.

12. Angevaren M, Aufdemkampe G, Verhaar H, Aleman A, Vanhees L. Physical activity and enhanced fitness to improve cognitive function in older people without known cognitive impairment. Cochrane Database Syst Rev 2008;3:CD005381.

13. Cheval B, Darrous L, Choi K, et al. Physical activity and general cognitive functioning: A Mendelian Randomization study. bioRxiv 2020. https://doi.org/10.1101/2020.10.16.342675.

14. Colcombe S, Kramer AF. Fitness effects on the cognitive function of older adults: a meta-analytic study. Psychol Sci 2003;14:125–130.

15. Mok A, Khaw K-T, Luben R, Wareham N, Brage S. Physical activity trajectories and mortality: population based cohort study. BMJ 2019;365.

16. Elhakeem A, Murray ET, Cooper R, Kuh D, Whincup P, Hardy R. Leisure-time physical activity across adulthood and biomarkers of cardiovascular disease at age 60–64: a prospective cohort study. Atherosclerosis 2018;269:279–287.

17. Jefferis BJ, Whincup PH, Lennon LT, Papacosta O, Goya Wannamethee S. Physical activity in older men: Longitudinal associations with inflammatory and hemostatic biomarkers, N-terminal pro-brain natriuretic peptide, and onset of coronary heart disease and mortality. J Am Geriatr Soc 2014;62:599–606.

18. Wannamethee SG, Shaper AG, Walker M. Changes in physical activity, mortality, and incidence of coronary heart disease in older men. Lancet 1998;351:1603–1608.

19. Almeida OP, Khan KM, Hankey GJ, Yeap BB, Golledge J, Flicker L. 150 minutes of vigorous physical activity per week predicts survival and successful ageing: a population-based 11-year longitudinal study of 12 201 older Australian men. Br J Sports Med 2014;48:220–225.

20. Gregg EW, Cauley JA, Stone K, et al. Relationship of changes in physical activity and mortality among older women. JAMA 2003;289:2379–2386.

21. Stessman J, Hammerman-Rozenberg R, Cohen A, Ein-Mor E, Jacobs JM. Physical activity, function, and longevity among the very old. Arch Intern Med 2009;169:1476–1483.

22. Aggio D, Papachristou E, Papacosta O, et al. Trajectories of physical activity from midlife to old age and associations with subsequent cardiovascular disease and all-cause mortality. J Epidemiol Community Health 2020;74:130–136.

23. Laddu D, Parimi N, Cauley JA, et al. The association between trajectories of physical activity and all-cause and cause-specific mortality. J Gerontol A Biol Sci Med Sci 2018;73:1708–1713.

24. Saint-Maurice PF, Coughlan D, Kelly SP, et al. Association of leisure-time physical activity across the adult life course with all-cause and cause-specific mortality. JAMA network open 2019;2:e190355.

25. Sanchez-Sanchez JL, Izquierdo M, Carnicero-Carreño JA, García-García FJ, Rodríguez-Mañas L. Physical activity trajectories, mortality, hospitalization, and disability in the Toledo Study of Healthy Aging. J Cachexia Sarcopenia Muscle 2020;11:1007–1017.

26. Nagin DS. Group-based trajectory modeling: an overview. Handbook of quantitative criminology 2010:53–67.

27. Nagin DS, Odgers CL. Group-based trajectory modeling in clinical research. Annu Rev Clin Psychol 2010;6:109–138.

28. Formánek T, Csajbók Z, Wolfová K, et al. Trajectories of depressive symptoms and associated patterns of cognitive decline. Sci Rep 2020;10:1–11.

29. Barnett TA, Gauvin L, Craig CL, Katzmarzyk PT. Distinct trajectories of leisure time physical activity and predictors of trajectory class membership: a 22 year cohort study. Int J Behav Nutr Phys Act 2008;5:1–8.

30. Laddu DR, Cawthon PM, Parimi N, et al. Trajectories of the relationships of physical activity with body composition changes in older men: the MrOS study. BMC geriatrics 2017;17:1–10.

31. Laddu DR, Parimi N, Stone KL, et al. Physical Activity Trajectories and Associated Changes in Physical Performance in Older Men: The MrOS Study. J Gerontol A Biol Sci Med Sci 2020;75:1967–1973.

32. Börsch-Supan A, Brandt M, Hunkler C, et al. Data resource profile: the Survey of Health, Ageing and Retirement in Europe (SHARE). Int J Epidemiol 2013;42:992–1001.

33. Rosen WG. Verbal fluency in aging and dementia. J Clin Exp Neuropsychol 1980;2:135–146.

34. Harris S, Dowson J. Recall of a 10-word list in the assessment of dementia in the elderly. Br J Psychiatry 1982;141:524–527.

35. Bauman AE, Reis RS, Sallis JF, et al. Correlates of physical activity: why are some people physically active and others not? Lancet 2012;380:258–271.

36. Cheval B, Maltagliati S, Sieber S, et al. Why are individuals with diabetes less active? The mediating role of physical, emotional, and cognitive factors. Ann Behav Med 2021:kaaa120.

37. Choi KW, Chen C-Y, Stein MB, et al. Assessment of bidirectional relationships between physical activity and depression among adults: a 2-sample mendelian randomization study. JAMA Psychiat 2019;76:399–408.

38. Kirk MA, Rhodes RE. Occupation correlates of adults’ participation in leisure-time physical activity: a systematic review. Am J Prev Med 2011;40:476–485.

39. Trost SG, Owen N, Bauman AE, Sallis JF, Brown W. Correlates of adults’ participation in physical activity: review and update. Med Sci Sports Exerc 2002;34:1996–2001.

40. United Nations Educational. International Standard Classification of Education 1997. UNESCO, Paris 2006.

41. Prince MJ, Reischies F, Beekman AT, et al. Development of the EURO–D scale–a European Union initiative to compare symptoms of depression in 14 European centres. Br J Psychiatry 1999;174:330–338.

42. Jung T, Wickrama KA. An introduction to latent class growth analysis and growth mixture modeling. Soc Personal Psychol Compass 2008;2:302–317.

43. Van De Schoot R, Sijbrandij M, Winter SD, Depaoli S, Vermunt JK. The GRoLTS-checklist: guidelines for reporting on latent trajectory studies. Struct Equ Modeling 2017;24:451–467.

44. Nguyen HQ, Herting JR, Kohen R, et al. Recreational physical activity in postmenopausal women is stable over 8 years of follow-up. J Phys Act Health 2013;10:656–668.

45. Lounassalo I, Salin K, Kankaanpää A, et al. Distinct trajectories of physical activity and related factors during the life course in the general population: a systematic review. BMC Public Health 2019;19:1–12.

46. Sabia S, Dugravot A, Dartigues J-F, et al. Physical activity, cognitive decline, and risk of dementia: 28 year follow-up of Whitehall II cohort study. BMJ 2017;357:j2709.

47. Daly M, McMinn D, Allan JL. A bidirectional relationship between physical activity and executive function in older adults. Front Hum Neurosci 2015;8:1044.

48. Sink KM, Espeland MA, Castro CM, et al. Effect of a 24-month physical activity intervention vs health education on cognitive outcomes in sedentary older adults: the LIFE randomized trial. JAMA 2015;314:781–790.

49. Snowden M, Steinman L, Mochan K, et al. Effect of exercise on cognitive performance in community-dwelling older adults: review of intervention trials and recommendations for public health practice and research. J Am Geriatr Soc 2011;59:704–716.

50. Young J, Angevaren M, Rusted J, Tabet N. Aerobic exercise to improve cognitive function in older people without known cognitive impairment. Cochrane Database Syst Rev 2015;4:CD005381.

51. Colzato LS, Kramer AF, Bherer L. Editorial special topic: enhancing brain and cognition via physical exercise. J Cogn Enhanc 2018;2:135–136.

52. Roig M, Nordbrandt S, Geertsen SS, Nielsen JB. The effects of cardiovascular exercise on human memory: a review with meta-analysis. Neurosci Biobehav Rev 2013;37:1645–1666.

53. Cotman CW, Berchtold NC. Exercise: a behavioral intervention to enhance brain health and plasticity. Trends Neurosci 2002;25:295–301.

54. Hillman CH, Erickson KI, Kramer AF. Be smart, exercise your heart: exercise effects on brain and cognition. Nat Rev Neurosci 2008;9:58–65.

55. Lisanne F, Hsu CL, Best JR, Barha CK, Liu-Ambrose T. Increased aerobic fitness is associated with cortical thickness in older adults with mild vascular cognitive impairment. J Cogn Enhanc 2018;2:157–169.

56. Cotman CW, Berchtold NC, Christie L-A. Exercise builds brain health: key roles of growth factor cascades and inflammation. Trends Neurosci 2007;30:464–472.

57. Raichlen DA, Alexander GE. Adaptive capacity: an evolutionary neuroscience model linking exercise, cognition, and brain health. Trends Neurosci 2017;40:408–421.

58. Frith E, Loprinzi P. Physical activity and individual cognitive function parameters: unique exercise-induced mechanisms. JCBPR 2018;7:92–106.

59. World Health Organization Global Health Observatory. Prevalence of insufficient physical activity. 2010. http://www.who.int/gho/ncd/risk_factors/physical_activity/en/. Accessed September 6, 2018.

60. Singh AS, Saliasi E, Van Den Berg V, et al. Effects of physical activity interventions on cognitive and academic performance in children and adolescents: a novel combination of a systematic review and recommendations from an expert panel. Br J Sports Med 2019;53:640–647.

61. Cheval B, Radel R, Neva JL, et al. Behavioral and neural evidence of the rewarding value of exercise behaviors: a systematic review. Sports Med 2018;48:1389–1404.

62. Cheval B, Rebar AL, Miller MM, et al. Cognitive resources moderate the adverse impact of poor neighborhood conditions on physical activity. Prev Med 2019;126:105741.

63. Cheval B, Tipura E, Burra N, et al. Avoiding sedentary behaviors requires more cortical resources than avoiding physical activity: an EEG study. Neuropsychologia 2018;119:68–80.

64. Cheval B, Daou M, Cabral DAR, et al. Higher inhibitory control is required to escape the innate attraction to effort minimization. Psychol Sport Exerc 2020;51:101781.

65. Cheval B, Boisgontier MP. The theory of effort minimization in physical activity (TEMPA). Exerc Sport Sci Rev in press.

